# USER-CENTERED DESIGN AND USABILITY EVALUATION OF A CANCER PREVENTION WEB APPLICATION: AN ITERATIVE APPROACH IN GERMANY

**DOI:** 10.1101/2025.04.29.25326679

**Authors:** Pricivel Carrera, Odile Elias, Ruidong Zhang, Tobias Norajitra, Angela Goncalves, Klaus Maier-Hein

## Abstract

**Objectives:** Limited public awareness of cancer risk factors necessitates effective dissemination of cancer prevention information. Digital technologies offer an opportunity to address this gap, yet there is scant information on tools for communicating cancer prevention evidence. This article describes the user-centered design, development and usability evaluation of a web application for personalized cancer prevention tailored to the German population.

**Materials and Methods:** Prototypes of the web-app were developed through early and continuous formative evaluations. These prototypes integrated validated cancer risk prediction models and recommendations using an evidence-based risk communication approach. In a graphical user interface (GUI) test usability was assessed using the system usability scale (SUS), deriving scores for overall usability, usefulness, and learnability. Qualitative data on user experience (UX) and user interface (UI) issues were also collected through think-aloud protocols, interviews, and questionnaires.

**Findings:** The GUI test showed a SUS score of 69.7/100 and a usefulness score of 75.8, indicating acceptable usability, while the learnability score was 48.4. Eight categories of UX/UI problems were identified, including one severe and three moderate issues related to data input, user guidance and risk visualization. Qualitative feedback highlighted strengths in navigation, information presentation, and interactive features such as the risk simulation tool.

**Discussion:** The iterative development and early user testing yielded valuable feedback, identifying key usability concerns during prototyping. The usability score was within an acceptable range, and the usefulness score was above average. However, the lower learnability score indicated potential challenges in user understanding and satisfaction. Identified usability issues highlight areas for improvement while positive feedback supports the design choices, particularly the use of visual aids, numerical data, and personalized feedback to improve risk comprehension and motivate behavior change.

**Conclusion:** The NCPC cancer prevention web application represents a significant step towards effective digital health promotion in Germany. Addressing identified usability concerns through continued iterative refinement and user involvement is crucial for enhancing the tool’s effectiveness. The integration of evidence-based risk communication strategies shows promise in improving risk comprehension and motivating behavior change among users.

## BACKGROUND AND SIGNIFICANCE

Cancer remains a significant global health challenge, with an estimated 30–50% of all cancer cases being preventable^1^. Metabolic, occupational, environmental, and behavioral factors have been implicated in 4.45 million deaths and 105 million disability-adjusted life-years (DALYs), accounting for 44.4% of cancer-related deaths and 42.0% of cancer DALYs in 2019^2^. An international survey revealed varying awareness of cancer risk factors across high-income countries, highlighting the need for better risk communication as increased knowledge of risk factors is positively associated with risk-reducing behaviors.^3^ Notably, countries with higher public knowledge of cancer risk factors have a greater proportion of individuals actively trying to reduce their personal cancer risk.^3^

Personalized digital technologies present a promising method for promoting cancer prevention. In particular, digital tools, such as personalized cancer risk calculators, have been developed for the general public in the United States^4^, Europe^5,6^, and Australia^7^. Most of these tools are tailored to specific cancer types^4,5^ and are intended to be used together with health professionals, as in the case of QCancer® developed for the English population^6^. The Cancer Risk Calculator developed for the Australian population offers prevention information based on Australian guidelines and cancer research, educating individuals on how their lifestyle choices can reduce their cancer risk^7^. However, since it does not provide an individual risk calculation of developing the disease it would be more accurate to describe this tool as a cancer prevention information guide or a lifestyle risk assessment tool^8^. In addition, there is limited information regarding the design and development of these tools to guide related efforts aimed at disseminating evidence and communicating recommendations for cancer prevention to the public^9^.

## OBJECTIVES

This paper aims to describe the user-centered and iterative development of a cancer prevention web-application prototype developed by the National Cancer Prevention Center (NCPC) in Germany, as well as to evaluate the usability of the high-fidelity prototype through a graphical user interface (GUI) test and presenting its results. In the context of software engineering, the usability of a software indicates the “extent to which a system, product or service can be used by specified users to achieve specified goals with effectiveness, efficiency and satisfaction in a specified context of use” as defined by the International Organization for Standardization (ISO)^10^.

## MATERIALS AND METHODS

To date, there is a lack of evidence-based, user-centered digital tools for primary and secondary cancer prevention specifically designed for the German population. In response, the NCPC is developing a web-app to disseminate cancer prevention information based on current evidence and clinical recommendations and to provide evidence-based personalized advice on primary cancer prevention and assessments of corresponding risk based on lifestyle data^11^. Additionally, this tool is designed to be effective, with the ultimate goal of achieving widespread adoption and regular use. The first part of this section provides an overview of the iterative design process and the functionality of the web-app. The second part details the GUI test and the methods used to collect and analyze usability data.

### Iterative development

The prototype was informed by several iterative formative evaluations during the initial stages of design and development. Starting in January 2022, consultations with stakeholders at the German Cancer Research Center (DKFZ) over five months shaped the foundational architectural design and mock-up creation for the web-app. Simultaneously, a review of evidence on population-based cancer prevention approaches^7^, including DKFZ’s contributions to primary and secondary prevention, was conducted^12,13^. Additionally, an analysis of relevant digital tools, identifying their respective pros and cons, informed the development of low-fidelity mockups^4–8^, which were used for visual exploration and communication of design ideas.

Early formative evaluations were integral to the iterative development process. Key informant interviews over two months provided feedback on the mock-ups. Subsequently, an interactive low-fidelity prototype with basic functionalities was developed over three months, followed by its formative evaluation involving focus groups with staff of the Prevention Outpatient Clinic, NCPC, and computer scientists between November 2022 and January 2023. Heuristic evaluations and cognitive walkthroughs were conducted on software applications associated with validated cancer risk assessment tools^14^.

These evaluations informed the design and development of the high-fidelity prototype, which will be described in subsequent sections. The findings led to refinements in content, functionality, and information architecture. Additionally, significant changes were made to the visual design, including the incorporation of risk visualization elements and modifications to the prototype layout.

### System architecture

The high-fidelity prototype comprises a frontend and a backend, as illustrated in Figure 1. The backend, developed in Python, utilizes Django as the framework and PostgreSQL as the database. It follows a microservice architecture and is deployed using Docker containers. The frontend uses Vue as the JavaScript framework and Vuetify for user interface (UI) components. The following section delves into the functionalities and features of the high-fidelity prototype.

**Figure 1.**
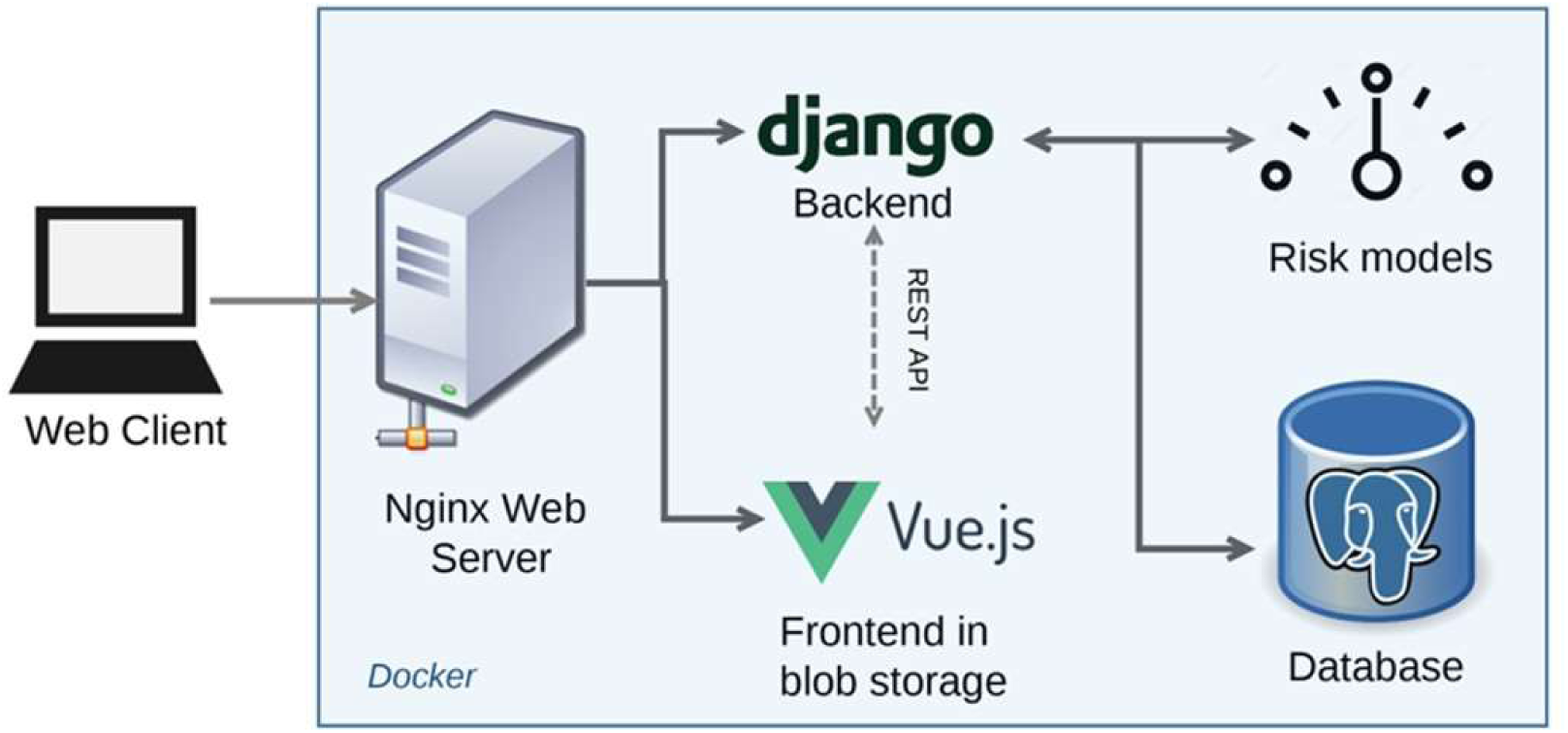
System Architecture of Germany’s National Cancer Prevention Center Web-app for Cancer Prevention.

### Multilayered web-app

To use the web-app, users first register by completing a multi-stage questionnaire, which includes questions about age, anthropometric measures, biological sex, physical activity, smoking history, alcohol consumption, diet and screening history. Figure 2 illustrates two stages of this questionnaire. Based on this information, a user profile is created, allowing for future updates if lifestyle changes occur.

**Figure 2.**
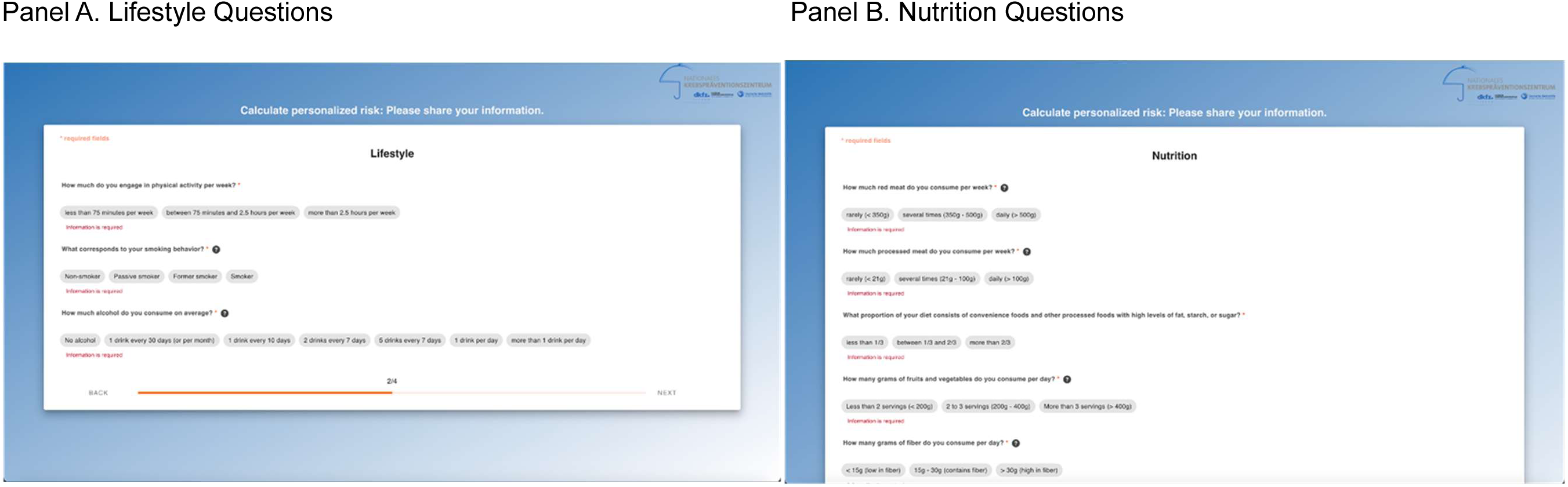
Sample Questions Used to Generate User Profiles and User Adherence to Cancer Prevention Recommendations.

Personalized feedback is provided based on the user’s profile. Following an implementation of the evidence-based cancer prevention recommendations by the World Cancer Research Fund/American Institute for Cancer Research (WCRF/AICR) in 2018^15^, the user’s profile data is leveraged to calculate their adherence level to lifestyle-based prevention recommendations. Compliance is categorized as strong, moderate, or poor. Additionally, the application offers evidence-based guidance, outlining advisable lifestyle adjustments and proposing preventive measures. This empowers users to understand their cancer risk and potentially mitigate it.

The dashboard, as shown in Figure 3, is the main interface. It displays the WCRF results and hosts various auxiliary modules. There, users can access additional informative primary prevention information, e.g. linking to in-person prevention clinic and a number of helpful interactive features, which are outlined briefly below.

**Figure 3.**
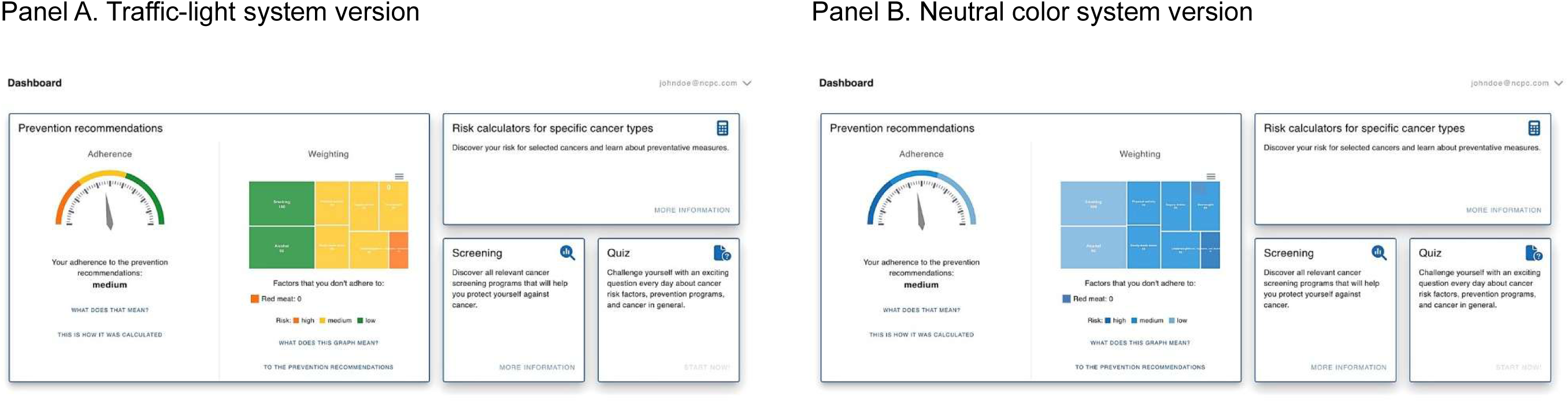
User Feedback on their Adherence to Cancer Prevention Recommendations.

The “screening” module presents an individual visual timeline of available population-based screening programs available in Germany, relevant to the user and ordered according to their schedule (see Figure 4). The timeline provides details on eligibility conditions, recommended age, year of eligibility for (or start of) screening, intervals, healthcare providers, and procedure descriptions^16^.

**Figure 4.**
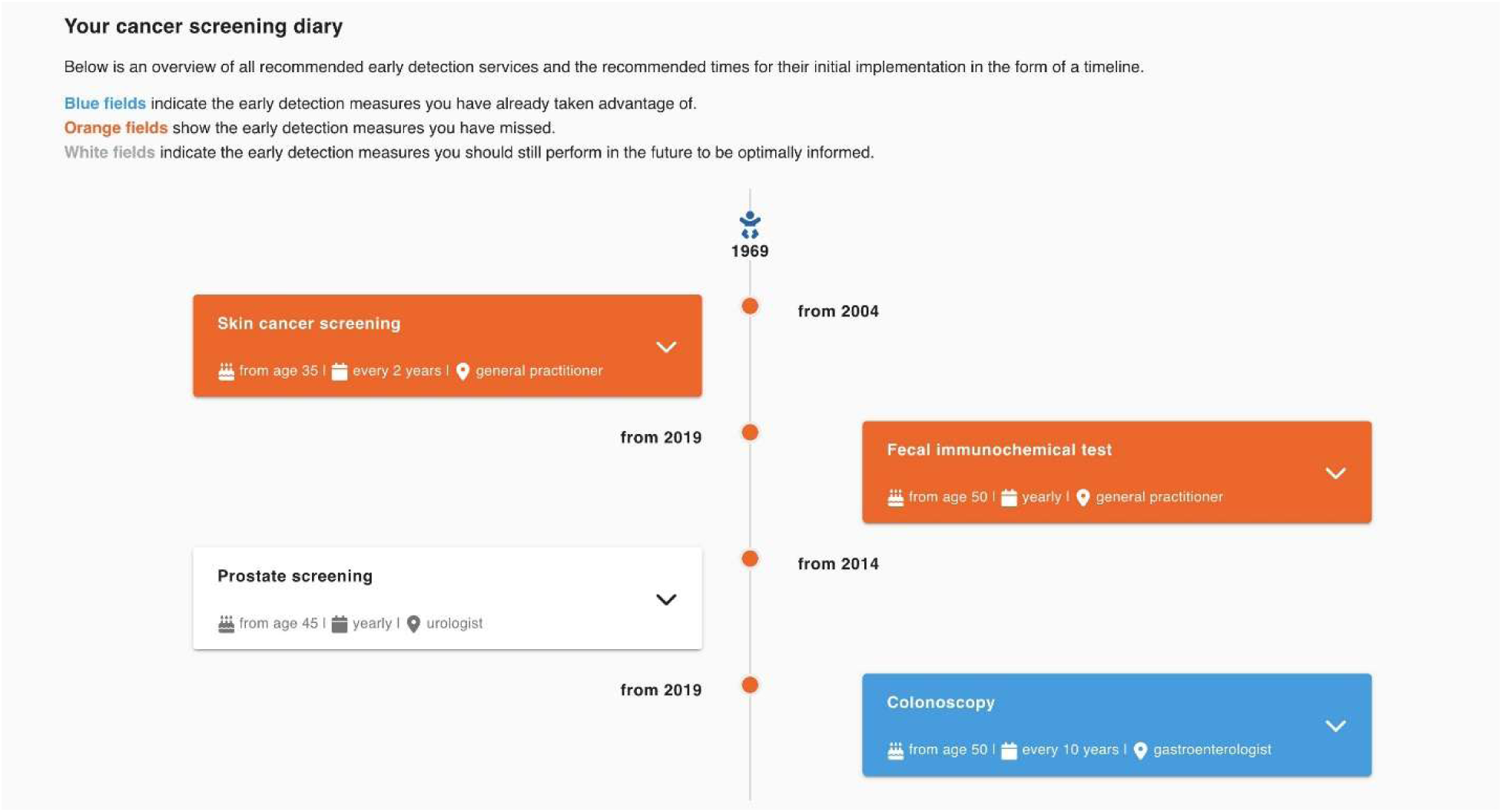
Personalized Cancer Screening Timeline.

Another key module is the calculation of specific cancer-type risks using eight validated lifestyle-based cancer risk models for the German population^12^. Additional lifestyle data not yet collected and used in the cancer-specific models are obtained via a specific questionnaire, complementing existing profile data. Results are output in categories of high, medium, or low risk, along with corresponding percentages (see Figure 5A). Moreover, users receive recommendations for lifestyle adjustments to reduce their personal risk, as illustrated in Figure 5B. Through a simulation tool, users can assess how lifestyle changes affect their risk, as shown in Figure 6, helping them create realistic improvement plans.

**Figure 5.**
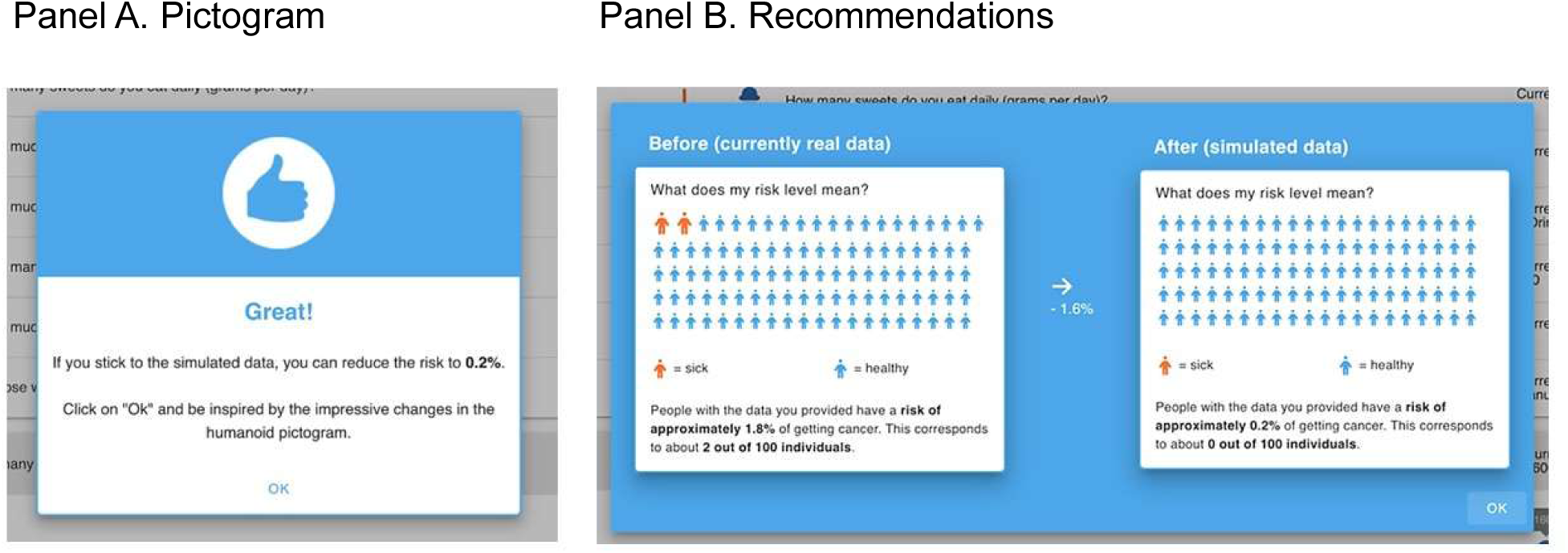
Communicating Colorectal Cancer Risk Using Pictogram and Recommendations for Risk-reduction.

**Figure 6.**
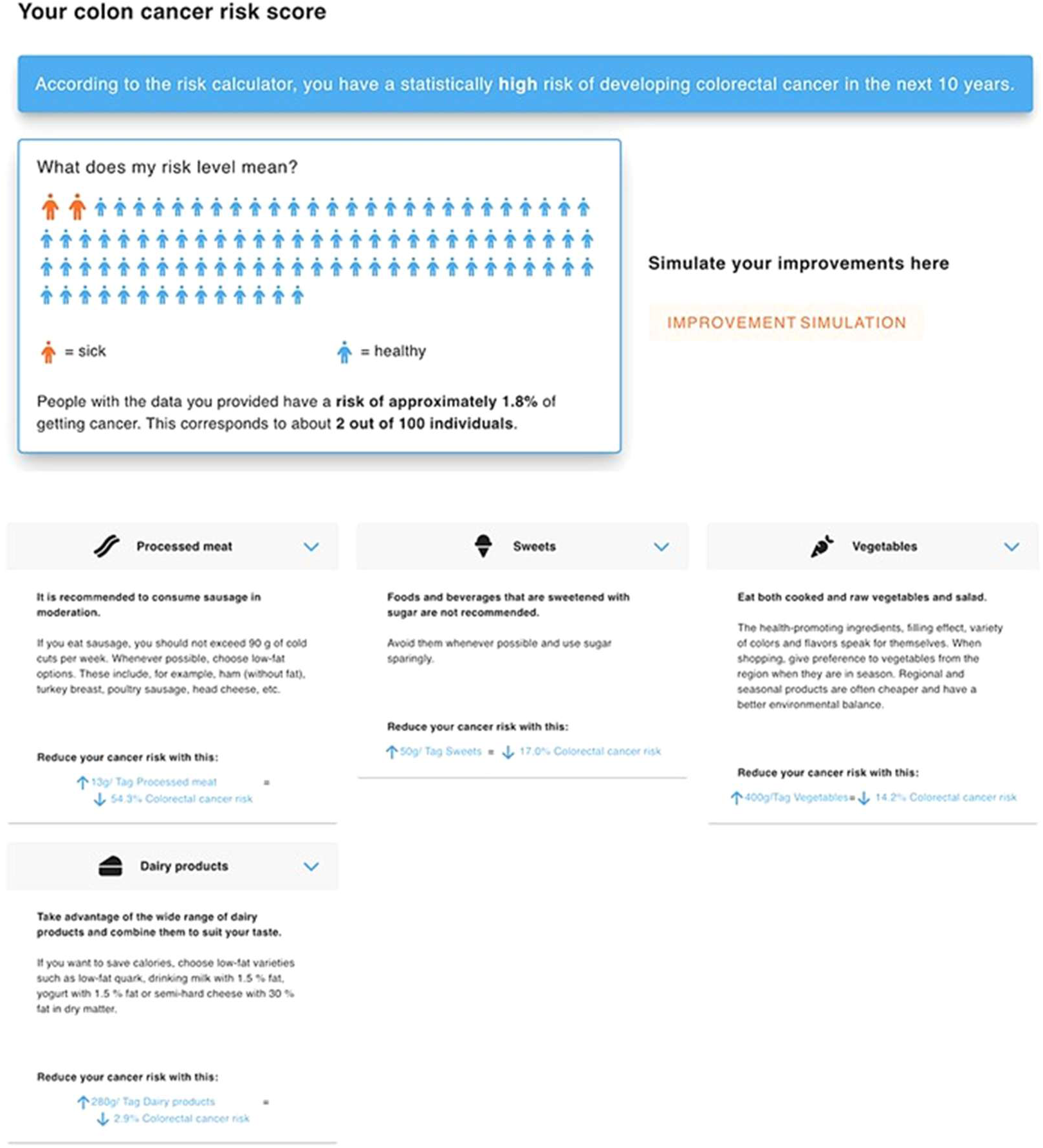
Colorectal Cancer Risk Simulation with Risk-modification and Results.

### Risk communication tools

Research shows that using multiple formats to communicate risk is more effective than using individual formats alone^17,18^. Therefore, the risk communication strategy combines both multiple visual and textual elements.

To visualize the adherence to the WCRF cancer prevention recommendations, two color schemes were implemented for categorical risk feedback. Users can switch between them according to their preferences. The default scheme uses the common traffic light system, shown in Figure 3A, which has been proven effective in influencing dietary decisions through the use of food labels. Notably, labels featuring a red indicator have been shown to encourage healthier dietary choices^19,20^. For inclusivity and accessibility, the color for the negative signal was adjusted to red-orange instead of red. Given potential emotional reaction to cancer risk, a neutral color scheme, seen in Figure 3B, was also introduced to assess emotional responses during GUI testing. This scheme uses three shades of blue, with the darkest shade representing the most negative interpretation.

These color schemes were applied to both of the diagrams: the speedometer and treemap^21^. The speedometer display employs a dial gauge that informs the user intuitively about their overall adherence level to the WCRF cancer prevention recommendations across three categories: low (red-orange), medium (yellow-orange), and high (green) (see Figure 3).

Additionally, a treemap helps users understand their adherence to each cancer risk factor, showing the weight assigned to each factor and enables simultaneous comparisons among multiple risk factors (see Figure 3)^22^. The color schemes were also applied to visualize the adherence to each risk factor, combined with smileys indicating the level of adherence. The use of smileys was informed by recent studies that demonstrated their effectiveness in indicating positive or negative signals^23^.

Figure 3 illustrates user feedback on their adherence to WCRF cancer prevention recommendations using the speedometer and treemap combined within the dashboard interface. It shows the application of the traffic light system (see Figure 3A) and the alternative neutral color scheme (see Figure 3B).

An additional level of risk communication included displaying specific risk numbers alongside the textual risk level to provide users with a more precise understanding of their risk level. Pictograms serve as simplified graphic symbols to convey complex information like cancer risk, transcending literacy barriers and appealing to diverse demographic groups ^24^. This enhances transparency and understanding of risk levels. The pictograms are used for communicating the risk for specific cancer types (see Figure 4) and visualizing the impact of simulated lifestyle changes on specific cancer risks (see Figure 5), motivating users to make positive lifestyle modifications for their health.

### User testing

Graphic user interface testing aims to evaluate the usability, functionality, and overall user experience of a software application’s graphical interface.^25^ In our research, the goal was to ensure that the interface elements, such as buttons, menus, icons, and visual components, function as intended and provide a seamless interaction for the end user.

A pilot test was conducted on August 1, 2023, with a purposeful sample of four individuals from the DKFZ and the NCPC. The pilot aimed to evaluate and refine the GUI test procedures and instruments, focusing on task comprehension, questionnaire clarity and digital completion of pre- and post-test questionnaires.

### Test setup

From a selected sample of 17 DKFZ employees, nine individuals participated in the GUI test. According to Turner et al., seven participants are considered sufficient to identify 90% of usability problems^26^. To evaluate the usability of all app pages and functions, eight tasks were developed to guide participants through the application. These tasks were provided sequentially to participants and were to be completed within a predefined time-frame. Each participant was assigned a persona to embody during the testing process.

Three personas were created to represent different user groups, improving result comparability and enhancing understanding of results based on the background provided for each persona^27–28^. The descriptions of these personas are provided in the Annex.

The test session began with a 10-minute introduction, where participants consented to screen recording and test participation and complete a pre-test questionnaire. Participants then had 45 minutes to perform the tasks while screen and voice recordings were conducted. The session concluded with a 5-minute outro, including a post-test questionnaire and an open interview for qualitative feedback on their experience using the web-app.

### Data acquired

Both quantitative and qualitative data were collected. Quantitative data included performance time and task scores from the test phase as well as responses from digital questionnaires conducted using the platform “Questionstar”^29^.

The pre-test-questionnaire gathered participants’ demographic information, motivation to use the web-app, and expectations about the cancer prevention web-app. The post-test questionnaire employed the SUS, a widely recognized and commonly used metric for measuring software usability^30^. It consists of ten items rated on a 5-point Likert scale, providing an overall usability assessment based on ISO 9241-11 standards, which cover effectiveness, efficiency, and satisfaction. Scores range from 0 to 100, with scores above 68 indicating above-average usability. Adapting SUS’s two-dimensional structure^31,32^, additional scores for “Usefulness” (including Utility, Complexity, Simplicity, Integration, Unity, Convenience, and Satisfaction) and

“Learnability” (evaluating Technical Support and Prior Learning) were derived. To standardize these scores for comparability, multiplication factors of 12.5 to learnability and 3.125 to usefulness^33^ were applied, yielding three individual metrics.

Additionally, the net promoter score (NPS) was calculated based on the last post-test questionnaire question, reflecting overall customer satisfaction and loyalty. Although not specific to UI design or health information system evaluation, NPS reflects user’s general sentiment toward a product or service^34^. Participants rated their likelihood of recommending the web-app to a friend or colleague on a scale of 0 (Not at all likely) to 10 (Very likely). Qualitative data were collected through “loud thinking” during tests, recorded via screen and voice capture, open interviews conducted after the test sessions, and free-text answers from the post-test questionnaire.

### Data analysis

To minimize bias, data analysis involved independent review of datasets and recordings, along with triangulation across findings, methods and investigators^35^. PC, OE and RZ independently analyzed recordings of each session. PC and RZ independently scored each participant’s performance on the tasks and completion times, while PC and OE independently evaluated the pre- and post-test-questionnaires. Overall mean scores for Usability, Usefulness and Learnability were calculated and analyzed, along with individual SUS item scores ^36^ and the NPS score. Data analysis was performed using Microsoft Office Excel 2019.

## RESULTS

Nine GUI test sessions were conducted to evaluate the usability of the high-fidelity prototype of a web-app for cancer prevention. However, one participant had difficulty impersonating the assigned persona, potentially introducing bias into the results. Consequently, data from this participant were excluded from the analysis to ensure comparability and validity, leaving data from eight participants for analysis. Among these participants, six were female and two were male, with an average age of 36 years (ranging from 33 to 58 years).

### Quantitative results

Table 1 gives an overview of task performance scores and time taken by each participant during the test.

**Table 1.**
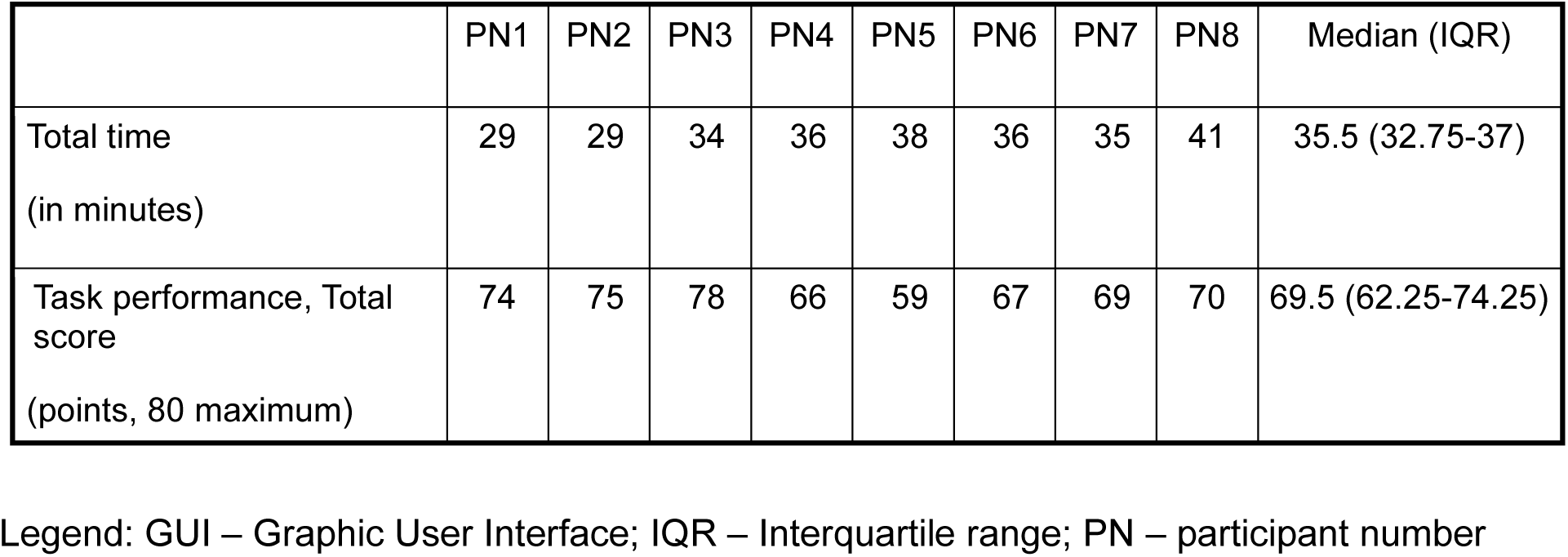
Task Duration and Performance of participants during the GUI Test.

Table 2 presents the SUS items, along with the average and median scores from all participants, as well as Usability, Learnability and Usefulness scores. Additionally, the NPS score was -25.

**Table 2.**
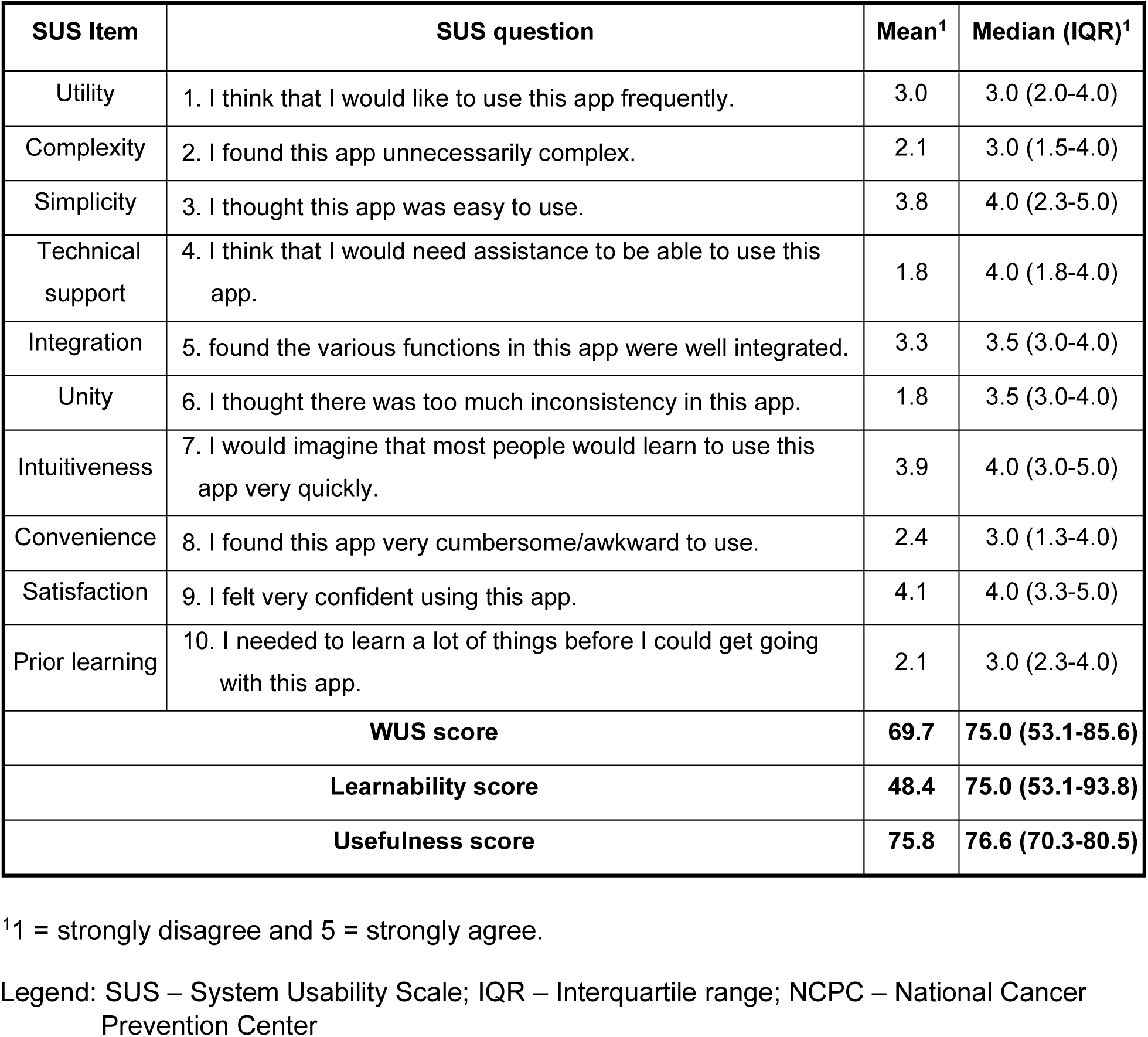
Web-app Usability Scale (WUS), Learnability and Usability Scores of the High-fidelity Prototype of NCPC Web-app. ^1^

### Qualitative Findings

Qualitative insights were gathered from the analysis of video recordings, the post-test questionnaires, interviews and observer notes. Eight categories of GUI problems were identified and classified by severity, detailed in Table 3. High severity problems pose a risk to core functionality, data integrity, or security. Moderate problems can create significant suboptimal experiences and workflow, but can be navigated by the user. These issues cover UX problems such as language clarity, explanations and guidance and the usability of scales and answer options. Low severity issues do not threaten functionality but hinder fluent app usage, including problems such as information presentation, navigation clarity and error handling. Findings addressed both UX/UI aspects, namely risk visualization, feedback mechanisms and information presentation.

**Table 3.** Usability Problems Identified and Their Severity.

| UX/UI problem category | Problem severity |
| --- | --- |
| 1. Inability to input data | High |
| 2. Scales and answer options not user-friendly | Moderate |
| 3. Unclear risk visualization and unmotivating feedback | Moderate |
| 4. Unclear or ambiguous explanation and guidance | Moderate |
| 5. Complex or technical language | Low |
| 6. Unappealing and lengthy presentation of information | Low |
| 7. Errors | Low |
| 8. Navigation elements not visible and unclear | Low |

Legend: UX – User experience; UI – User interface

Although several UX/UI problems were identified, participants praised specific design elements and functions. Notably, there was unanimous preference for the traffic-light scheme with either the speedometer or treemap over the neutral color scheme. Additionally, users universally preferred information presentation in square tiles, particularly for cancer risk factors. Positive feedback was received for tooltips in questionnaires, aiding comprehension of questions and providing guidance on nutritional information. The effective use of icons throughout the app to convey information at a glance was also appreciated. Five users praised the recommended lifestyle actions for reducing cancer risk, highlighting the informative target value and concise summaries provided. Four users expressed strong interest on using the simulation tool to modify cancer risk through lifestyle changes. They also found the exclamation mark effective in highlighting the most impactful risk factors for risk reduction. Six users appreciated the clarity and organization of information provided for early screening on the screening timeline.

## DISCUSSION

The GUI test results provide valuable insights into the usability of the cancer prevention webapp prototype. The overall Usability score of 69.7 falls within the acceptable range, indicating that the prototype meets basic usability requirements^25–27^. Research suggests that scores below 68 indicate design issues requiring further research, while scores exceeding 68 indicate the need for minor design refinements^37^. Compared to home healthcare devices, the prototype scored higher than the EpiPen and pregnancy test but lower than the inhaler or thermometer^38^. With a higher Usefulness score of 75.8, participants perceived the prototype as beneficial and potentially valuable for cancer prevention efforts. However, the lower Learnability score of 48.4 suggests that users may have faced challenges in understanding and navigating the prototype, potentially requiring additional guidance or training.

The NPS of -25 indicates the need for improvements in user satisfaction. Nonetheless, it’s important to note the limitations of NPS, especially its simplicity, which may not fully capture the complexities of user satisfaction in healthcare settings when using solely ^39,40, 41,42^. Despite these shortcomings, integrating NPS insights with our findings on usability, usefulness, and learnability offers a more comprehensive understanding of the user experience.

Matching the quantitative scores, the qualitative findings led to minor adjustments in the content, functionality, and architecture of the web-app, along with significant alterations to the visual design. Key changes included integrating risk visualization elements and modifying the prototype layout.

A primary concern was the clarity of data input, requiring more intuitive layout elements. Participants were confused by the sequence of entering information, especially regarding waist circumference and waistband options, as well as the units for inputting data. This confusion hindered progress in completing the questionnaire. Similar issues arose with differing start and end values in answer options, particularly linked to problem 2. In the simulation tool, users expected dynamic risk updated with each input change, rather than only seeing changes after clicking to compile their altered lifestyle plan. To address these issues, users recommended a standardized layout for data entry, alongside the adoption step-by-step window tutorials or comprehensive video tutorials to enhance user guidance.

The integration of tooltips to explain individual factors or questionnaire terms was well received by participants, although some terms remained unclear. While the simulation aimed for universal exploration regardless of risk level, it allowed actions potentially increasing risk. An exclamation mark indicated the most impactful lifestyle factors to reduce risk, which participants found beneficial as orientation. However, low-risk users especially were confused due to the lack of specific cues on optimal lifestyle behavior for risk reduction. This was particularly notable when encountering unexpected information, such as the risk reduction associated with increased dairy consumption for colorectal cancer prevention.

Despite content clarity not being the primary focus of the GUI test, these results highlight the need for additional explanations, expanded tooltips with examples of optimal food and lifestyle values, and further elucidation of specific risk visualizations to address problems 2 and 3 and to enhance the learnability of the functionalities^43^. Recommendations to address minor issues include simplifying explanatory text and incorporating more imagery and symbols, particularly on risk factor information pages, to ensure broad comprehension among the German population, especially given the fast-paced, stressful nature of everyday life.

Positive findings highlighted several notable strengths in the app’s design and functionality. Participants praised the navigation system and menu layout for easy access to various application sections. Icons across the interface improved user comprehension and expedited information assimilation. The structured layout and concise information provided in the screening timeline aided users in maintaining an overview of their screening actions understanding the existence of the screenings, and recognizing their importance. Participants valued the tooltips for offering helpful contextual information and guidance. Although they suggested including better examples, these tooltips helped individuals from diverse backgrounds understand the prevention information and specific recommendations. Notably, the adoption of a traffic light schema for grading and risk communication offered clear visual cues for users to intuitively interpret their risk levels. Specifically in visualizing adherence to specific risk factors, the use of smileys underscored the level’s meaning and helped users quickly absorb information about their status.

The implementation of these visual elements, and their combined use, enhanced understanding of evidence-based risk and adherence levels in multi-level risk communication. Consequently, the acceptance of visual elements, aligned with expectations and existing research, remained valuable for informing our design choices and prototype development. Especially the combined use of multiple visual and textual elements supports inclusivity by providing alternative ways to convey information. Moreover, the feedback from participants clearly indicated an interest in using the web-app and its interactive modules. These positive findings underscore the effectiveness of these design decisions in enhancing the usability of the web-app.

## CONCLUSION

The iterative development process yielded valuable user feedback during the prototyping phase of the NCPC cancer prevention web-app. GUI test results indicated that the prototype could be improved to enhance user experience. Identified problems emphasize the need for refinements, particularly in addressing critical issues such as data input, user-friendly elements and the implementation of a tutorial system. Positive participant feedback, especially regarding the simulator and traffic light system, supported our design choices and the use of evidence-based risk communication tools to enhance risk comprehension and motivate behavior change. These findings underscore the importance of such a digital tool in disseminating evidence-based cancer prevention information and promoting healthy behaviors in the German population. By addressing the usability concerns, the NCPC cancer prevention web-app represents a significant step towards effective health promotion and cancer prevention.

## OUTLOOK

After resolving the identified issues, a new version of the web-app will undergo usability testing with potential end-users as part of the iterative development process. This testing phase will assess the alpha version of the web-app, aligning with the ongoing development of the high-fidelity prototype.

## Author Contributions

PC, AG and KMH designed the study. PC, RZ, AG, and KMH conceptualized the web-app. PC, OE, RZ, AG, and KMH designed the web-app. PC, OE and RZ executed the GUI test and analyzed the data generated. RZ developed the backend of the web-app and cancer risk models. OE implemented the user interface and frontend of the web-app. PC drafted the manuscript and all the other co-authors revised it critically. All authors reviewed the results and approved the final version of the manuscript.

## Acknowledgements

The authors would like to acknowledge the active collaboration and valuable contribution of the following individuals in the design of the NCPC web-app, particularly in providing constructive feedback in early iterations of the NCPC web-app: Dr. Ursula Will of the NCPC Prevention Outpatient Clinic (POC); Dr Susanne Weg-Remers of the Cancer Information Service (KID); Prof. Karen Steindorf and Dr. Florian Herbolsheimer of the DKFZ Division of Physical Activity, Prevention and Cancer; Prof. Michael Hoffmeister of the DKFZ Division of Clinical Epidemiology and Aging Research; Drs Katharina Gudd and Romana Sowade, and Drs Anja Braun and Delia Braun of the DKFZ Strategic Planning Unit. The DCP would like to acknowledge the hosting of the pilot test and all user testing sessions of the NCPC web-app at the POC. Likewise, the DCP would like to acknowledge the invaluable contribution of participants to the GUI test and its pilot. This work was completed while PC and ZH were at the DKFZ.

## Annex. Profiles of the Three User Personas Developed for the Graphic User Interface Test of Web-app for Cancer Prevention Prototype

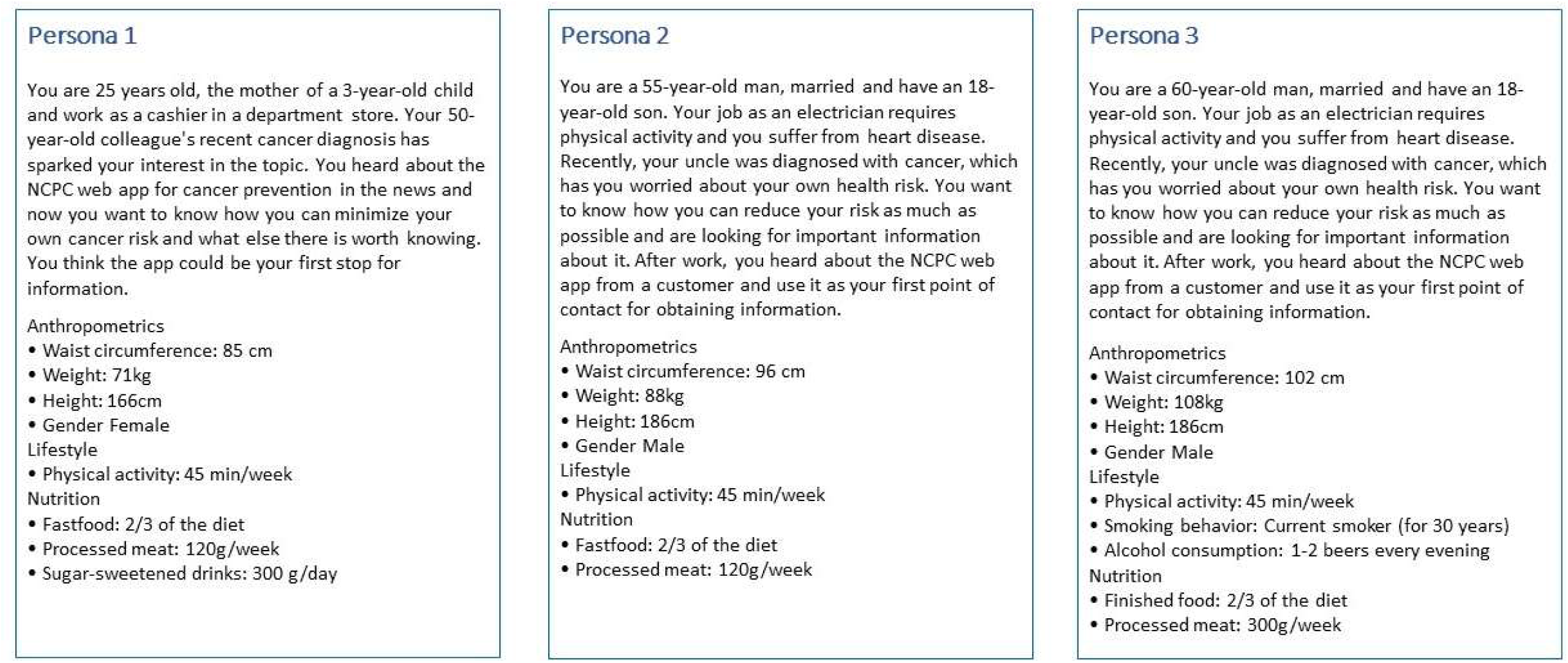

## Data availability

The data underlying this article are available in the article and in its online supplementary material.

